# COVID-19 vaccine hesitancy among persons living in homeless shelters in France

**DOI:** 10.1101/2021.04.29.21256256

**Authors:** C Longchamps, S Ducarroz, L Crouzet, N. Vignier, L. Pourtau, C Allaire, AC Colleville, T El Aarbaoui, M Melchior, the ECHO study group

## Abstract

COVID-19 vaccine hesitancy is frequent and can constitute a barrier to the dissemination of vaccines once they are available. Unequal access to vaccines may also contribute to socioeconomic inequalities with regard to COVID-19. We studied vaccine hesitancy among persons living in homeless shelters in France between May and June 2020 (n=235). Overall, 40.9% of study participants reported vaccine hesitancy, which is comparable to general population trends in France. In multivariate regression models, factors associated with vaccine hesitancy are: being a woman (OR=2.55; 95% CI 1.40-4.74), living with a partner (OR=2.48, 95% CI 1.17-5.41), no legal residence in France (OR=0.51, 95% CI 0.27-0.92), and health literacy (OR=0.38, 95% CI 0.21, 0.68). Our results suggest that trends in vaccine hesitancy and associated factors are similar among homeless persons as in the general population. Dissemination of information on vaccine risks and benefits needs to be adapted to persons who experience severe disadvantage.

## Introduction

As the COVID-19 pandemic spreads in many parts of the world, access to an effective vaccine offers hope of a new much-needed preventive measure in upcoming months. However, in the area of COVID-19 infection as well as with regard to other infectious diseases, there are concerns that vaccine hesitancy may be a barrier to rapid roll-out of a new vaccine. Indeed, in Europe, it is estimated that approximately one person out of four would not want to be vaccinated (1-3), young persons, women, and those not feeling at risk with COVID-19 being most likely to be reluctant. Among reasons cited to explain vaccine hesitancy, fear of side effects is the most frequent one (55% of vaccine hesitant individuals), particularly among women (3). In recent months, and particularly since the approval of several COVID-19 vaccines, opinion polls suggest that vaccine hesitancy has decreased. However, inequalities with regard to vaccination intentions remain (4).

Members of disadvantaged populations, appear disproportionately impacted by the COVID-19 epidemic. In particular, persons who live in shared facilities such as homeless shelters have been shown to be at high risk of getting infected (5). Moreover, migrants from low or middle-income countries have also been shown to have high levels of COVID-19 morbidity and mortality (6), and for those who live in homeless shelters the risks may be compounded. Elevated levels of COVID-19 morbidity and mortality among persons who experience socioeconomic disadvantage and migrants likely reflect a high prevalence of risk factors of severe COVID-19 infection (ex. diabetes, hypertension) and insufficient information access to the health care system (7). However there is also suggestion of lower levels of adherence to preventive measures, although data on this topic are lacking (8).

Persons residing in homeless shelters and migrants may constitute relevant target groups for COVID-19 vaccination, however vaccine acceptability in this population is not known. We studied the intention to be vaccinated against COVID-19 among persons living in homeless shelters in France, a majority of whom are recent migrants, integrating risk factors identified by the working group on vaccine hesitancy established by the WHO (9). The study was conducted in the Spring 2020, before anti-COVID-19 vaccines were approved for use.

## Methods

### Study population

Our investigation is based upon data collected in ECHO, a cross-sectional study conducted among residents of 18 short and long-term homeless shelters in the Paris (n=12), Lyon (n=5) and in Strasbourg (n=1) regions. The study interview, aiming to measure participants’ level of information regarding COVID-19, their knowledge, implementation and acceptability of various preventive measures, was conducted between May 2^nd^ and June 28, 2020, corresponding to the 1^st^ COVID-19 lockdown period (March 16-May 11, 2020) and its immediate aftermath, by trained interviewers, in person or by telephone. The questionnaire was administered in French, in English or in the participant’s language: 25% of questionnaires were completed with the assistance of a trained translator contacted by telephone (most frequently used languages: Arabic, Pashto, Dari, Tigrinya, and Amharic). In total, 929 residents were invited to take part in the survey, 131 refused participation, 263 were unavailable, and 535 (57.6%) completed the study questionnaire. An item on the intention to be vaccinated against COVID-19 was added to the ECHO questionnaire from the end of May 2020, and 235 study participants provided information.

The ECHO study received approval of the Ethical Research Committee of the University of Paris (CER-2020-41).

## Measures

The intention to be vaccinated was ascertained by the following item: “If a vaccine existed would you be willing to get vaccinated”? Participants who responded ‘no’ or ‘I don’t know’ were considered probably vaccine hesitant.

Factors potentially associated with ‘vaccine hesitancy’, which we studied, based on the SAGE consortium, included sociodemographic characteristics (sex, age, household composition, presence of children, WHO region of birth, administrative status, educational level, French language level, employment prior to lockdown, health insurance, duration of residence in the homeless shelter, social support), health (depression ascertained with the PHQ-9 using a cut-off of 10 to identify persons with a significant level of symptoms (10), and self-reported chronic health problems, fear of getting infected by COVID-19), as well as health-seeking information (trust in official information about COVID-19, health literacy ascertained using the ‘*Appraisal of health information’* subscale (n=5 items) of the Health Literacy Questionnaire (HLQ)(11), primary sources of COVID-19 information).

### Statistical analysis

To examine factors associated with the intention to be vaccinated against COVID-19, we conducted bivariate analyses with the chi-square statistic. Next all variables associated with the study outcome with a p≤0.25 were included in a step-wise logistic regression model.

All statistical analyses were conducted using the R software version 1.3.1093.

## Results

As shown in **Table 1**, ECHO study participants were mainly male (66.3%), young (61.1% below 35 years of age), lived alone and without children (respectively 81.5 and 65.5%), were migrant (92.5% born outside of France), did not have legal residence in France (60.8%), had low educational attainment (62.9% with secondary school education or less), low or intermediate level knowledge of the French language (80.7%), no employment prior to the COVID-19 lockdown (70.5%), and resided in the shelter where they were interviewed for a short period of time (60.4% at most one year). In terms of health, 27.9% of participants had symptoms of clinical depression, 25.5% reported another chronic disease, 67.0% feared getting infected by COVID-19, 76.4% expressed trust in official information on COVID-19, 49.3% had low health literacy, and the leading sources for information about COVID-19 were the TV (63.0%) and social media (84.0%).

**Table 1.**
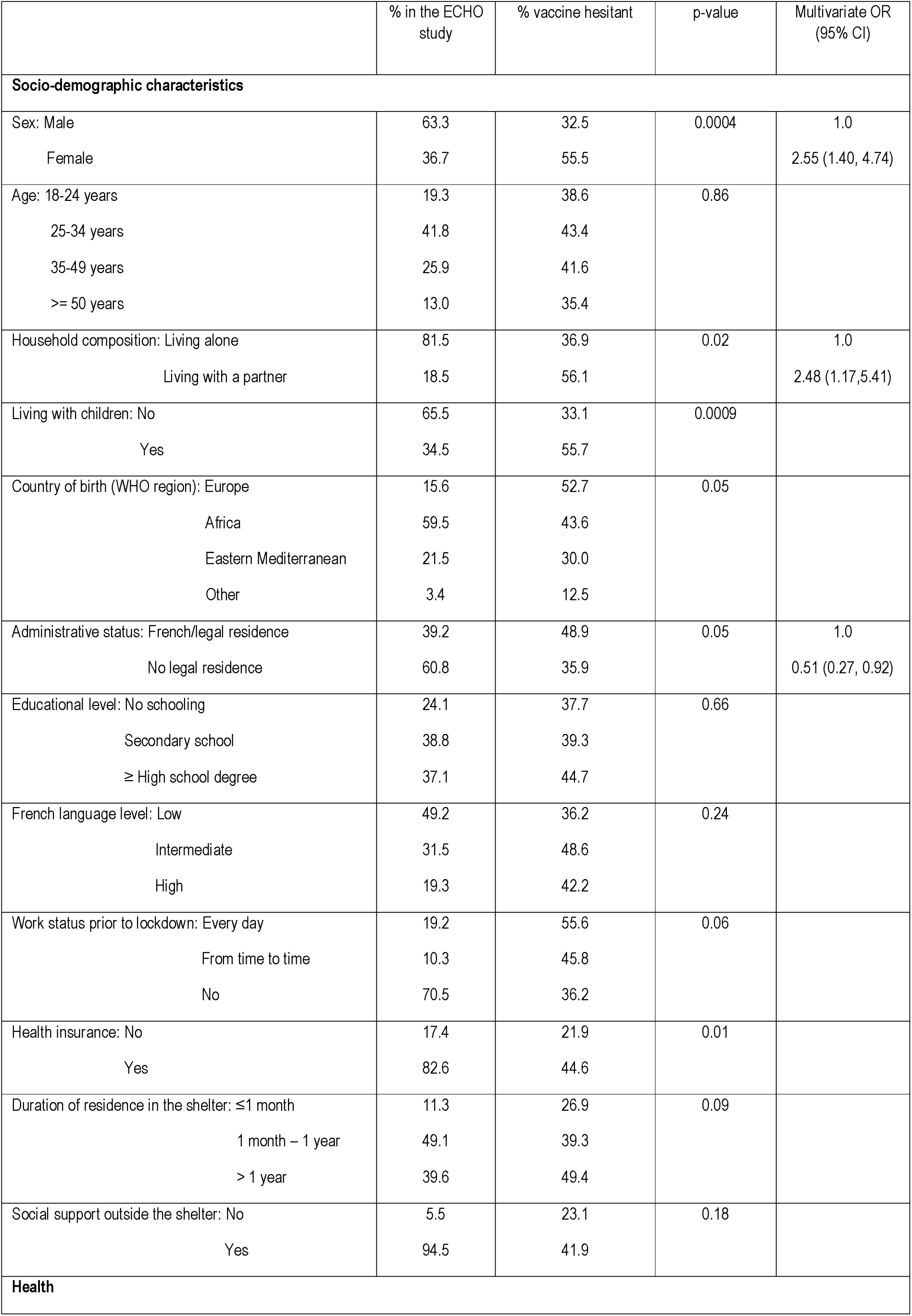
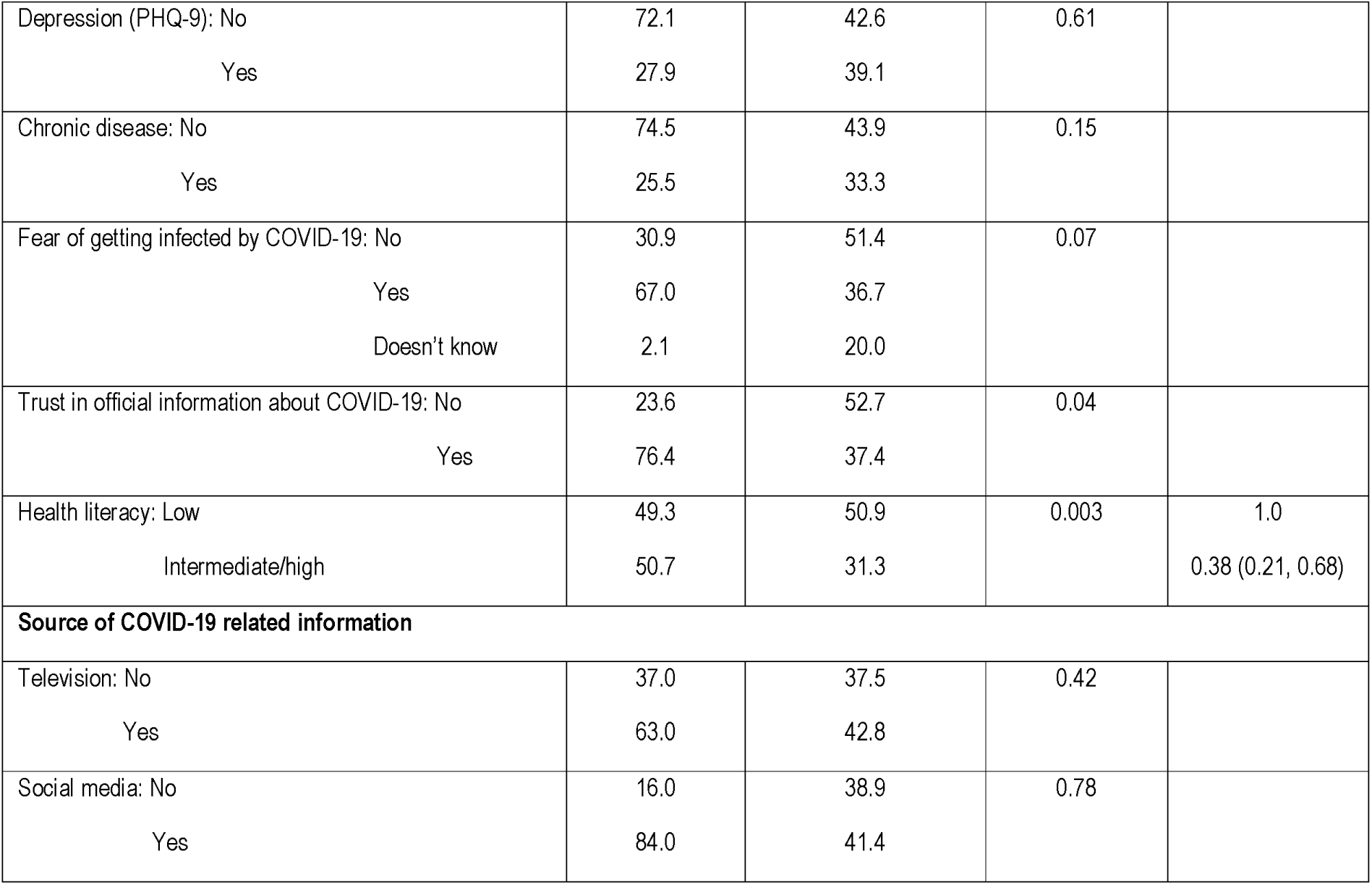
Factors associated with COVID-19 vaccine hesitancy (ECHO study, France, n=240, May-June, 2020, %, chi-square, multivariate logistic regression, OR; 95% CI)

Overall, 40.9% reported not being willing to get vaccinated against SARS-CoV-2 – among those 71.2% indicated they would not want to be vaccinated and 28.8% did not know what to answer-. In bivariate analyses factors associated with ‘vaccine hesitancy’ included sex (55.9% in women vs. 32.5% in men, p=0.0004), household composition (living with a partner: 56.1 vs. living alone: 36.9%, p=0.0232), living with children (yes: 55.7 vs. no: 33.1%, p=0.0009), region of birth (Europe: 52.7%, Africa: 43.6%, Eastern Mediterranean: 30.0%, Other: 12.5%, p=0.05), legal status (French/legal residence: 48.9% vs. no legal residence: 35.9%, p=0.05), and health literacy (low: 50.9% vs. intermediate/high: 31.3%, p=0.0026). In multivariate logistic regression models, factors which remained associated with ‘vaccine hesitancy’ were sex (female vs. male: OR=2.55; 95% CI 1.40-4.74), household composition (participants living with a partner vs. living alone: OR=2.48, 95% CI 1.17-5.41), administrative status (no legal residence vs. French/with legal residence: OR=0.51, 95% CI 0.27-0.92), and health literacy (intermediate/high vs. low: OR=0.38, 95% CI 0.21, 0.68).

## Discussion

Our study conducted among persons living in short and long-term homeless shelters in France, among whom a majority were migrant, shows approximately 41% of COVID-19 hesitancy in the Spring of 2020. Factors associated with low intention to be vaccinated against COVID-19 include female sex, living with a partner, as well as French citizenship/legal residence, and low health literacy. Our data suggest that factors associated with ‘vaccine hesitancy’ appear similar in this highly disadvantaged group to those reported in the general population, however their absolute contribution may be higher.

First, we need to address limitations and strengths of this study, which can influence data interpretation. First, ECHO is not representative of persons living in homeless shelters in France. However, we collaborated with several different organizations and surveyed 18 homeless shelters hosting different types of populations. Importantly, homeless shelters and participants were recruited independently of the risk of COVID-19 infection, which rules out the possibility of systematic bias. Moreover, our sample’s characteristics (a majority of young men who are migrant) is similar to that observed across homeless facilities across France (12). Second, we had no information regarding participants’ COVID-19 infection, which could influence perceptions of preventive measures and particularly vaccine acceptance. However at the time we conducted our study (May-June 2020), neither virological nor serological COVID-19 tests were easily accessible, making it difficult to ascertain the actual prevalence of infection. This lead us to focus on perceptions of the illness and its impact. Main strengths of ECHO are: a) the inclusion of a large sample of individuals living in homeless shelters in the two largest metropolitan areas of France (Paris and Lyon); b) access to translation services which made it possible to interview persons who did not speak French.

Our findings are generally in line with observations from general population surveys. Previous studies had suggested that women are more likely to express COVID-19 vaccine hesitancy than men (2, 3). In particular, they seem especially concerned about possible side effects, which may reflect overall higher levels of concerns about health (3). Additionally, women are more likely than men to seek information about health on the Internet and social media (13), which may put them in contact with negative information regarding vaccines. This suggests that it is necessary to better control information that is disseminated via the Internet, as well as provide persons with accurate information in these much accessed media.

Our study found that persons who live with a partner and are French or have legal residence in France are especially likely to report a low intention to be vaccinated against COVID-19. This finding may be related to a higher dissemination of anti-vaccine messages in media used by this group. It may also be that these persons have come across vaccine-critical content (in mainstream news media for instance) in the years prior to the COVID-19 epidemic, as controversies in this area have emerged in France as early as in 2009. Additionally, health care is not always adapted to migrants’ needs, resulting in inadequate or negative experiences, which they are likely to encounter with time (14). Prior studies reported an influence of political views on vaccine hesitancy (1), which recent migrants may be less exposed to. Moreover, we found a relationship between low health literacy, which indicates individuals’ limited capacity to seek and evaluate health information, and vaccine hesitancy. This is consistent with research on vaccine hesitancy in other areas (15) and indicates that all messages on measures which can prevent COVID-19 should be adapted to be easily understood by all.

In conclusion, our study shows elevated levels of low intention to be vaccinated against COVID-19 among persons living in homeless shelters, who may be at higher risk of COVID-19 infection. As this population is likely to be a relevant target group for future COVID-19 vaccine deployment (16), it is important to ensure that the information regarding the COVID-19 infection and vaccine is disseminated in an appropriate way and all barriers lifted, in order to inform properly and increase vaccine adherence.

## Data Availability

Data referred to in the manuscript are available from the Principal Investigators upon request.

